# Remote exercise-induced sweat chloride measurements using a wearable microfluidic sticker in cystic fibrosis patients

**DOI:** 10.1101/2025.03.05.25323327

**Authors:** Rachel S. Nelson, Margaret G. Grossman, Zasu M. Klug, Mia Calamari, Alvaro Donayre, Thaddeus Cybulski, Jared Schooley, Garett J. Griffith, Daniel M. Corcos, Donald E. Wright, Jessica C. Wallace, Da Som Yang, John A. Wright, John A. Rogers, Roozbeh Ghaffari, AJ Aranyosi, Manu Jain

**Affiliations:** Northwestern University Feinberg School of Medicine, Chicago, IL, USA; Northwestern University Division of Pulmonary and Critical Care Medicine, Chicago, IL, USA; Northwestern University School of Physical Therapy and Human Movement Sciences, Chicago, IL, USA; Epicore Biosystems, Inc. Cambridge, MA, USA; Chung-Ang University School of Mechanical Engineering, Seoul, South Korea; Querrey-Simpson Institute for Bioelectronics, Northwestern University Evanston, IL, USA; Northwestern, University Department of Materials Science and Engineering, Evanston, IL, 60202, USA; Department of Biomedical Engineering, Northwestern University, Evanston, IL, 60202, USA

## Abstract

Sweat parameters such as volume and chloride concentration may offer invaluable clinical insights for people with CF (PwCF). Pilocarpine-induced sweat collection for chloridometry measurement is the gold-standard for sweat chloride, but this technique is cumbersome and not suitable for remote settings. We have previously reported the utility of a skin-interfaced microfluidic device (CF Patch) in conjunction with a smartphone image processing platform that enables real-time measurement of sweating rates and sodium chloride loss in laboratory and remote settings. Here we conducted clinical studies characterizing the accuracy of the CF Patch compared to pilocarpine-induced sweat measurements using chloridometry and tested the feasibility of exercise-induced sweat chloride measurements in PwCF. The CF Patch demonstrated strong correlations compared to sweat chloride measured by chloridometry across clinic and remote settings and detected greater day-to-day sweat chloride variability in PwCF on CFTR modulators than healthy volunteers. These findings demonstrate that the CF Patch is suitable as a remote management device capable of measuring chloride concentrations and offers the potential of monitoring the efficacy of CF medication regimens.

## Introduction

Cystic fibrosis (CF) is a genetic disorder caused by mutations in the cystic fibrosis transmembrane conductance regulator (CFTR) gene, leading to impaired chloride (Cl^-^) transport across cell membranes.^1^ As a result, people with CF (PwCF) have higher Cl^-^ and sodium (Na^+^) concentrations in their sweat.^2^ The measurement of [Cl^-^] (chloride concentration) in sweat is a cornerstone in the diagnosis of CF, is associated with clinical outcomes^3^ and is also used as a biomarker in clinical trials.^4, 5^ In addition, sweat [Cl^-^] can serve as a tool to monitor patient health, assess adherence to CFTR modulators (small molecule therapies that restore CFTR protein function), and help adjust treatment regimens.^6^

Traditional methods of sweat collection for [Cl^-^] measurement involve the use of absorbent pads, gauzes, and rigid skin-interfacing cavities, like the Macroduct system (Macroduct Sweat Collection System; MSCS, ELITechGroup), followed by laboratory analysis with a chloridometer.^7^ These collection and post-analysis techniques employ sweat induction via the transdermal delivery of pilocarpine through iontophoresis.^8^ The multitude of steps and requirements involved in the pristine and reliable capture of eccrine sweat are limited to specialized referral centers, making remote sweat collection measurements challenging.

Recent advances in the development and deployment of wearable microfluidic biosensors have gained significant traction in remote care settings, with the promise to overcome the limitations of existing sweat collection strategies.^9^ In particular, the clinical utility of a microfluidic wearable platform was demonstrated in a feasibility study across pediatric and newborn CF patients compared to gold standard clinical techniques.^9^ Moreover, commercialization efforts have led to wearable microfluidic sweat patch solutions tailored for athletes,^10^ with smartphone mobile application software that enables real-time image processing of sweat loss and sodium chloride concentrations in remote settings.^11^ Taken together, this new class of wearable microfluidic technology facilitates non-invasive and exercise driven measurements of sweat [Cl^-^] and regional sweat rates during physical activity and exercise, with potential to help support adult PwCF outside of clinical settings.^11–13^

The present study aimed to compare and correlate sweat [Cl^-^] measurements obtained from a wearable CF sweat patch (CF Patch) to those from the gold standard MSCS and chloridometry. We also examined the relationship between [Cl^-^] in sweat produced by exercise and that induced by pilocarpine. Measurement of [Cl^-^] by the CF Patch during exercise at remote settings was used to assess the feasibility of measuring sweat [Cl^-^] remotely as well as to determine how well it correlates with sweat [Cl^-^] measured clinically. This wearable technology could revolutionize remote monitoring of PwCF, allowing for regular, accessible measurements of sweat chloride to assess several clinically useful parameters such as CFTR modulator adherence, pharmacodynamics and drug-drug interactions among others. In addition, since regular exercise is an important contributor to long-term health for PwCF,^14^ the CF Patch could also empower patients and caregivers with timely and actionable data for individualized hydration and electrolyte replenishment post-exercise, thus enhancing its utility.

## Results

### Soft wearable CF Patch design

The CF Patch consists of a soft and flexible polymeric material stack with an enclosed network of microfluidic channels, bioassay (corresponding to chloride concentrations), and food coloring dye (orange colored) embedded in microreservoirs. The CF Patch is manufactured using a roll-to-roll process, which enables multiple layers of soft polymers, embedded microchannels, patterned hypo-allergenic skin adhesive, and an ultrathin graphical capping layer to be constructed rapidly at speeds approaching 2,000 devices per minute. Figure 1 shows a schematic drawing of the CF Patch being worn on the inner volar forearm during physical exercise. Smartphone algorithms custom developed for image capture and processing enable real-time analysis of sweat loss, sweat rate, chloride concentration, and sodium chloride loss in near real-time following sweat capture upon completion of physical exercise.

**Figure 1.**
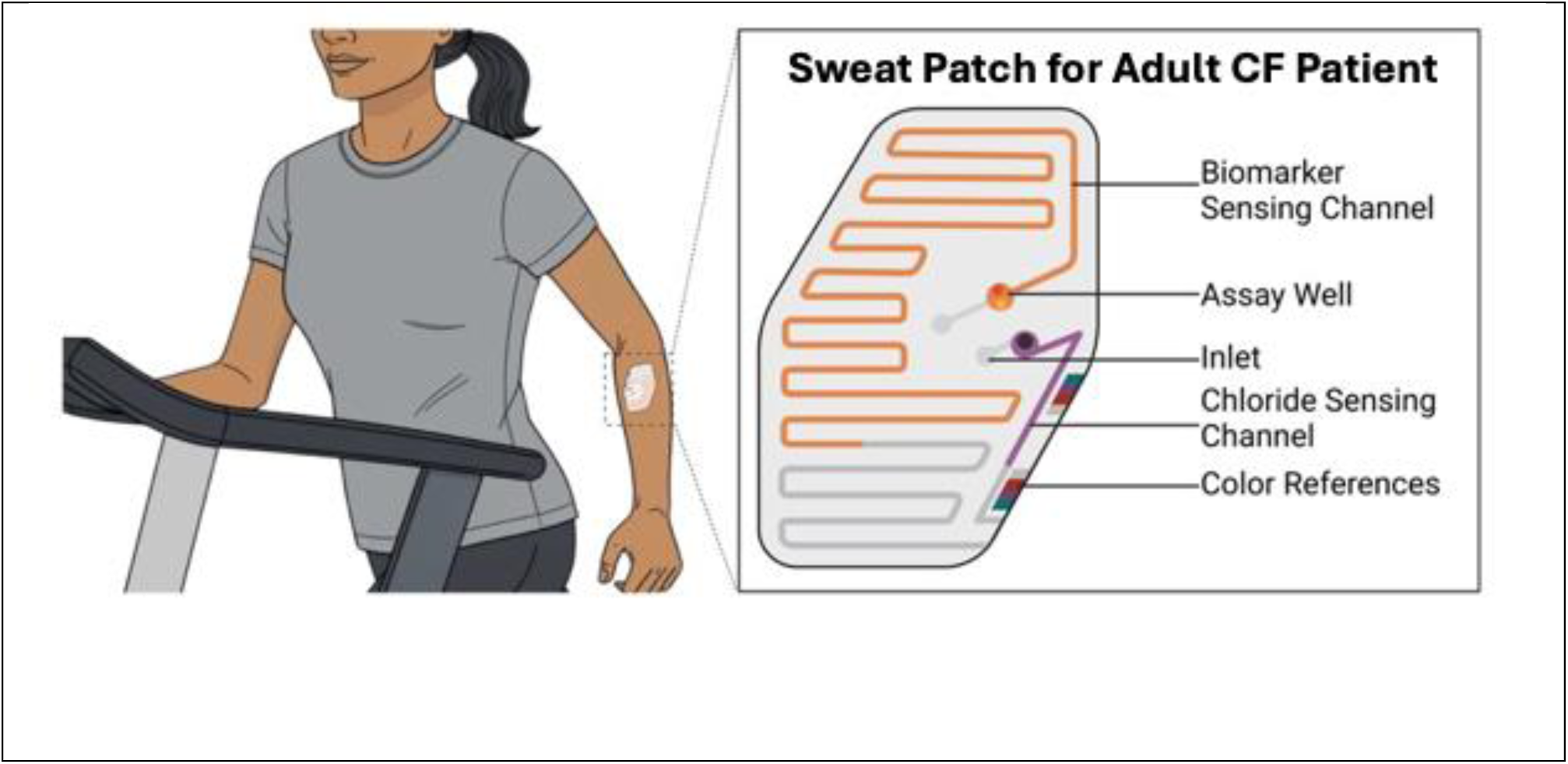
CF Patch. Schematic drawing of CF patient exercising on treadmill and wearing sweat sensor (CF Patch) on their volar forearm. The CF Patch contains two fluidic channels for: i) measuring regional and whole-body sweat loss (orange colored channel) and ii) chloride concentrations (purple colored channel). Color reference markers printed on the CF Patch enable image analysis in different lighting conditions.

### CF Patch sweat rate and loss analysis

The study combined multiple sweat collection and analysis protocols. An initial lab-based session consisted of three separate measurements:

- Pilocarpine-induced sweat collected by MSCS and analyzed with a chloridometer (“pilocarpine-MSCS”);
- Pilocarpine-induced sweat collected by the CF patch and analyzed colorimetrically (“pilocarpine-CF”);
- Exercise-induced sweat collected by the CF Patch and analyzed colorimetrically (“exercise-CF”).

In addition, a separate set of up to 5 home-based sessions using the exercise-CF protocol were performed.

Figure 2 shows a flow diagram of the sample size for each type of sweat collection. The final on-site sample size analyzed was *n* = 19 for pilocarpine-MSCS, *n* = 18 for pilocarpine-CF Patch, and *n* = 15 for on-site exercise-CF Patch. For the remote exercise part of the study, 18 PwCF completed all 5 exercise sessions and were able to successfully collect at least 1 sweat sample in the CF Patch. In addition, 15 PwCF met the pre-specified criteria of at least 2 successful CF Patch collections. All 7 healthy volunteers (HV) completed every phase of the study.

**Figure 2.**
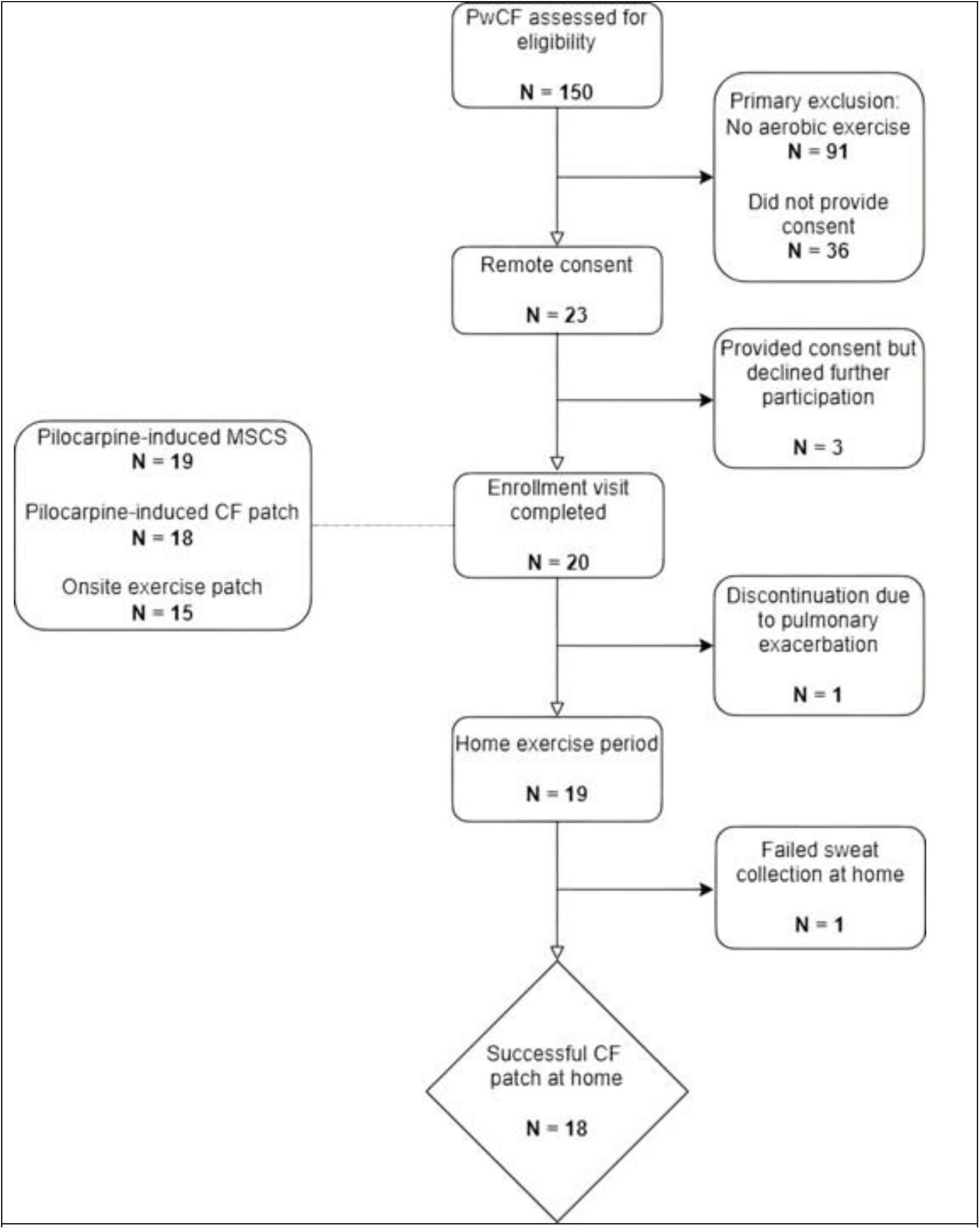
Flow diagram of study protocol.

Demographics and selected physiological measures show that PwCF and HV were well matched for the listed parameters (Table 1). The mean percent predicted forced expiratory volume in one second (ppFEV1) for PwCF was 93.3 ± 19%, and 18 of them were being treated with CFTR modulators. As measured by cardiopulmonary exercise testing (CPET) during the on-site visit, the peak oxygen uptake (VO_2peak_) was similar for HV and PwCF participants (34.4 ± 3.0 mL/kg/min and 30.9 ± 7.0 mL/kg/min, respectively). During the on-site constant wattage exercise session, during which sweat was collected in the CF Patch, the mean exercise time was 36.4 ± 3.8 minutes for HV and 40.5 ± 8.3 minutes for PwCF.

**Table 1.** Demographics of Enrolled Cohorts.

| Variable | PwCF<br>(n=20) | No CF (n=7) | p value |
| --- | --- | --- | --- |
| **Physiological Characteristics** |  |  |  |
| Age at enrollment, years<sup>*</sup> | 32.6 (20-64) | 30.7 (27-34) | 0.512 |
| Female (n, %) | 10 (55%) | 3 (42.9%) | 0.678 |
| Weight (lb)<sup>†</sup> | 162.9 (36.6) | 176.4 (43.7) | 0.481 |
| Height (in)<sup>†</sup> | 65 (14) | 68.2 (2.8) | 0.345 |
| Peak VO2 (mg/kg/min)<sup>†</sup> | 31.2 (6.9) | 34.4 (3) | 0.105 |
| **Ethnicity** |  |  |  |
| Non-Hispanic (%) | 100 | 100 | ns |
| **Race** |  |  |  |
| White (%) | 100 | 100 | ns |
| **Cystic Fibrosis medical history** |  |  |  |
| F508del homozygous | 10 (50%) |  |  |
| F508del heterozygous | 9 (45%) |  |  |
| Heterozygous, other | 1 (5%) |  |  |
| Diagnostic Sweat (mmol/L)<sup>‡,†</sup> | 93.3 (24.9) |  |  |
| FEV1 (L)<sup>†</sup> | 3.7 (1.1) |  |  |
| ppFEV1 (L)<sup>†</sup> | 94.9 (19.6) |  |  |
| Treated with CFTR modulators<sup>§</sup> | 19 (95%) |  |  |
| <sup>*</sup> Average and range<br><sup>†</sup> Average and standard deviation<br><sup>‡</sup> Two subjects did not have diagnostic sweat available.<br><sup>§</sup> The predominant CFTR modulator used in this cohort was ETI; additional modulators were Ivacaftor and open-label Vanzacaftor, which were documented in one participant each. |  |  |  |

### Comparison of MSCS chloridometry and CF Patch colorimetry

In order to validate the CF Patch for sweat [Cl^-^] measurement in PwCF, we first collected paired pilocarpine-induced sweat prior to exercise sweat collection at the on-site exercise visit. Pilocarpine collections for MSCS chloridometry and the CF Patch using colorimetry for [Cl^-^] measurements were successful in 19 and 18 PwCF respectively and both types of collections were successful in all 7 HV. PwCF had a mean sweat [Cl^-^] of 55 (range 17 – 106 mmol/L) by MSCS chloridometry and 66 (range 35 – 104 mmol/L) by the CF Patch. The results for HV were 25 mmol/L (range 10 – 49 mmol/L) by MSCS chloridometry and 29 mmol/L (range 5 – 54 mmol/L) by CF Patch colorimetry. There was a strong correlation between MSCS chloridometry and CF sweat patch derived chloride concentrations across the combined cohorts with an r=0.90 (*p*<0.0001) (Figure 3a). The Bland-Altman plot shows a mean bias of 12.7 mmol/L (CI: −11.9-37.3) without a systematic error as suggested by the confidence interval crossing 0 (Figure 3b). Thus, the CF sweat patch performed very well as a surrogate for MSCS chloridometry across a range of clinically relevant sweat chloride values and was able to detect CFTR modulator decreases in sweat chloride.

**Figure 3.**
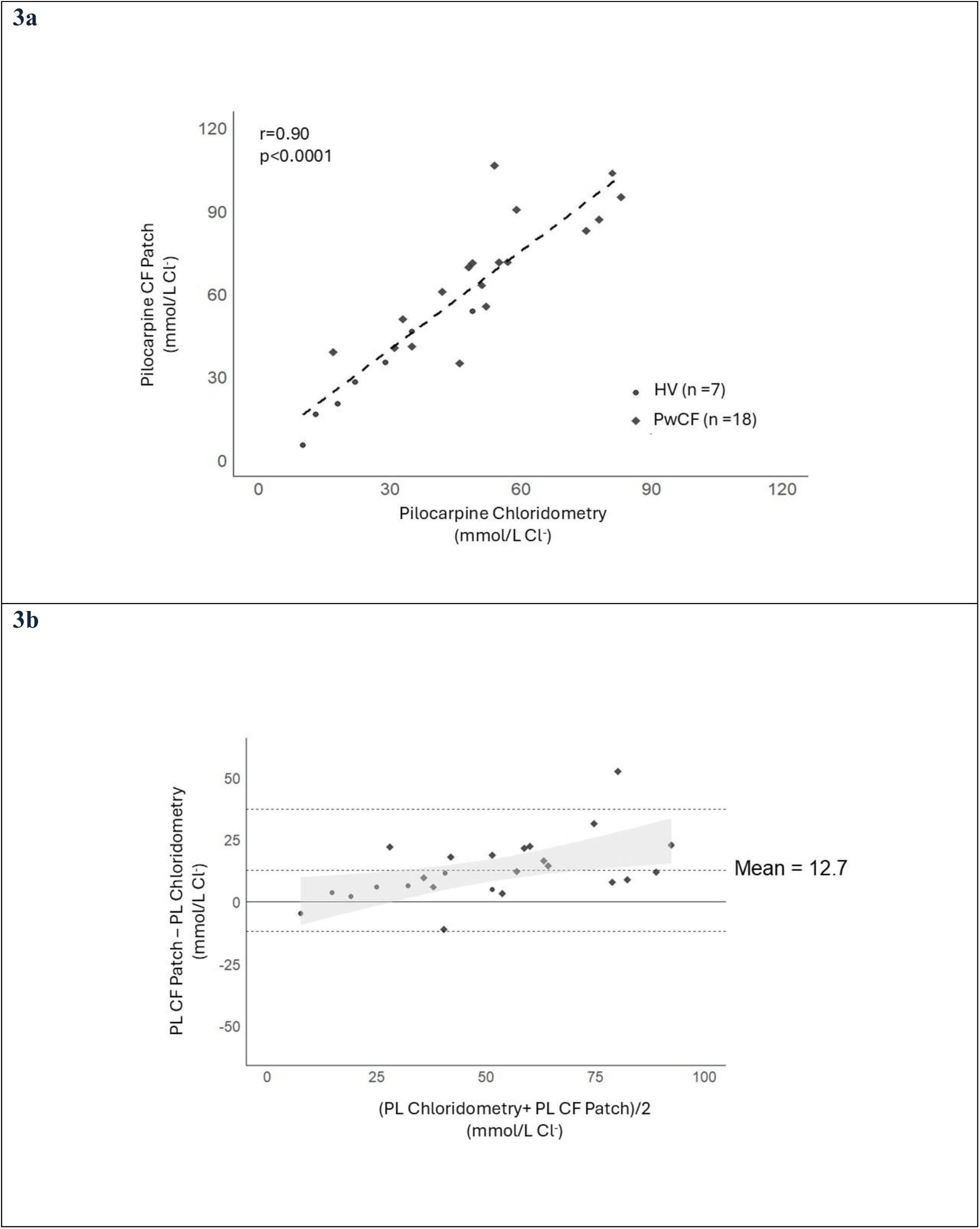
Pilocarpine-induced CF Patch sweat chlorides correlation versus MSCS chloridometry. People with CF and HV had resting paired pilocarpine induced sweat chlorides collected, one via a Macroduct for chloridometry and the other via a CF sweat patch. Shown is the regression line for the paired sweat chloride measurements for the combined CF and HV cohort (A). The Bland-Alman plot for assessing agreement between the CF sweat patch and chloridometry shows good agreement with limited bias. The dashed line represents the mean difference (bias), and the gray shaded area represent the limits of agreement (mean difference ± 1.96 standard deviations). The shaded area denotes the 95% confidence interval for the limits of agreement (B).

### Comparison of Pilocarpine and Exercise Laboratory Sweat Chlorides

Using the CF sweat patch affixed to each forearm during the exercise laboratory constant wattage session, we were able to collect sweat from 15 PwCF and 7 HV until adequacy of sample volume was visually confirmed. The mean sweat volume collected was 35 (+/-18 µl) and 43 (+/-16.6 µl) for PwCF and HV, respectively. For PwCF, the mean sweat chloride during exercise was 64 (range 18 – 96 mmol/L) and for HV it was 43 (range 8 – 79 mmol/L). There was a strong correlation between pilocarpine-induced and exercise-induced CF Patch sweat chloride concentrations with an r=0.79 (*p*<0.0001) (Figure 4a). The Bland-Altman plot showed a mean bias of 2.1 mmol/L (95% CI: −12.7.9-26.8) and no systematic error (Figure 4b). There was also an excellent correlation between pilocarpine-MSCS chloridometry and exercise CF Patch sweat chloride concentrations, r=0.81 (*p*< 0.0001) and no systematic bias (Figure 5a and 5b). This demonstrates that exercise can serve an adequate surrogate to pilocarpine for sweat induction and chloride measurement.

**Figure 4.**
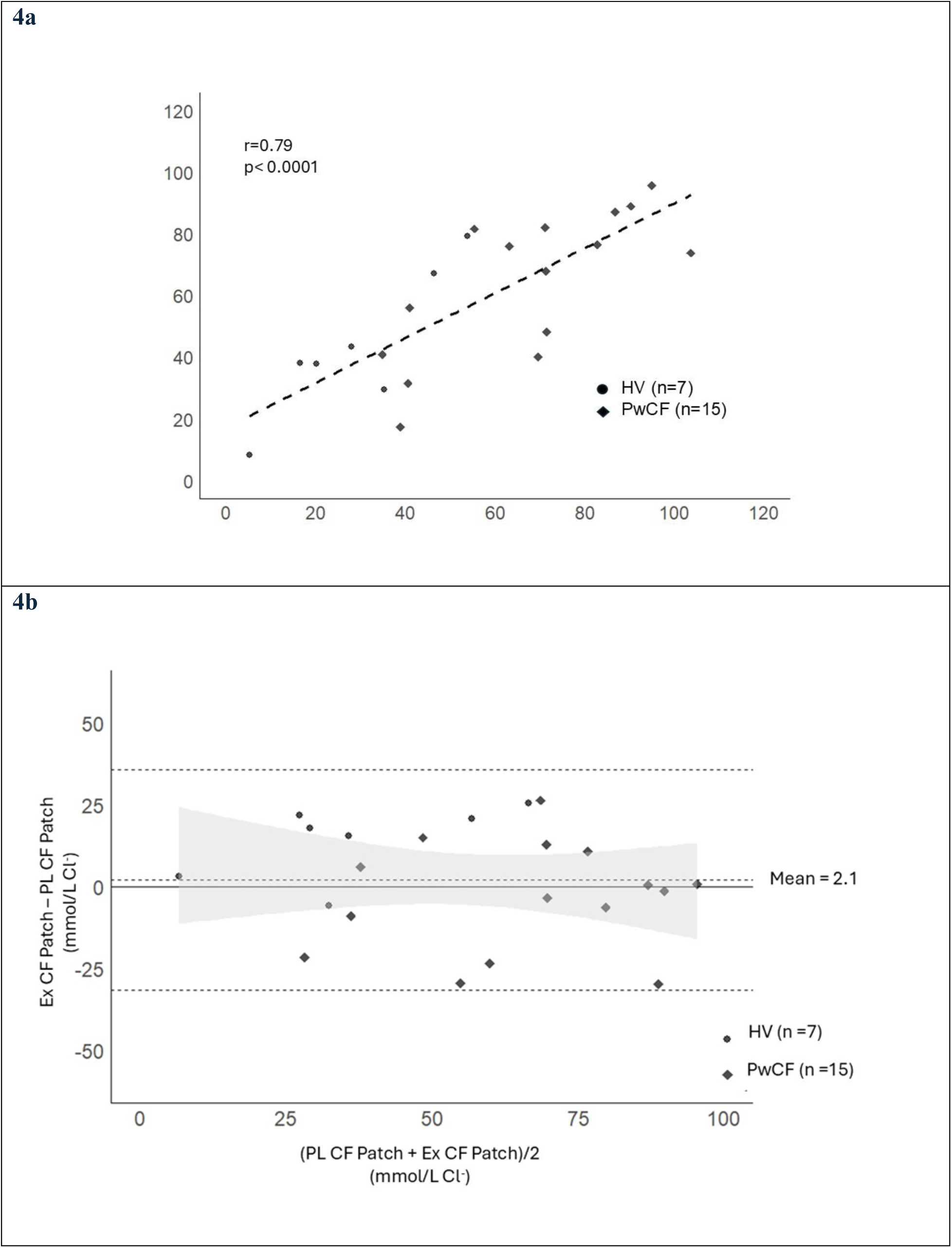
Exercise-induced CF Patch sweat chloride correlation versus pilocarpine-induced CF Patch sweat chloride. People with CF and HV had sweat collected with the CF patch during constant work exercise and it was compared to Pilocarpine induced sweat chloride measured by CF patch. Shown is the regression line for the comparison sweat chlorides between pilocarpine and exercise for the combined CF and HV cohort (a). The Bland-Alman plot for assessing agreement between the CF sweat patch and chloridometry shows good agreement and almost no bias. The dashed line shows the mean difference (bias), and the gray shaded area represents the limits of agreement (mean difference ± 1.96 standard deviations). The shaded area denotes the 95% confidence interval for the limits of agreement (b).

**Figure 5.**
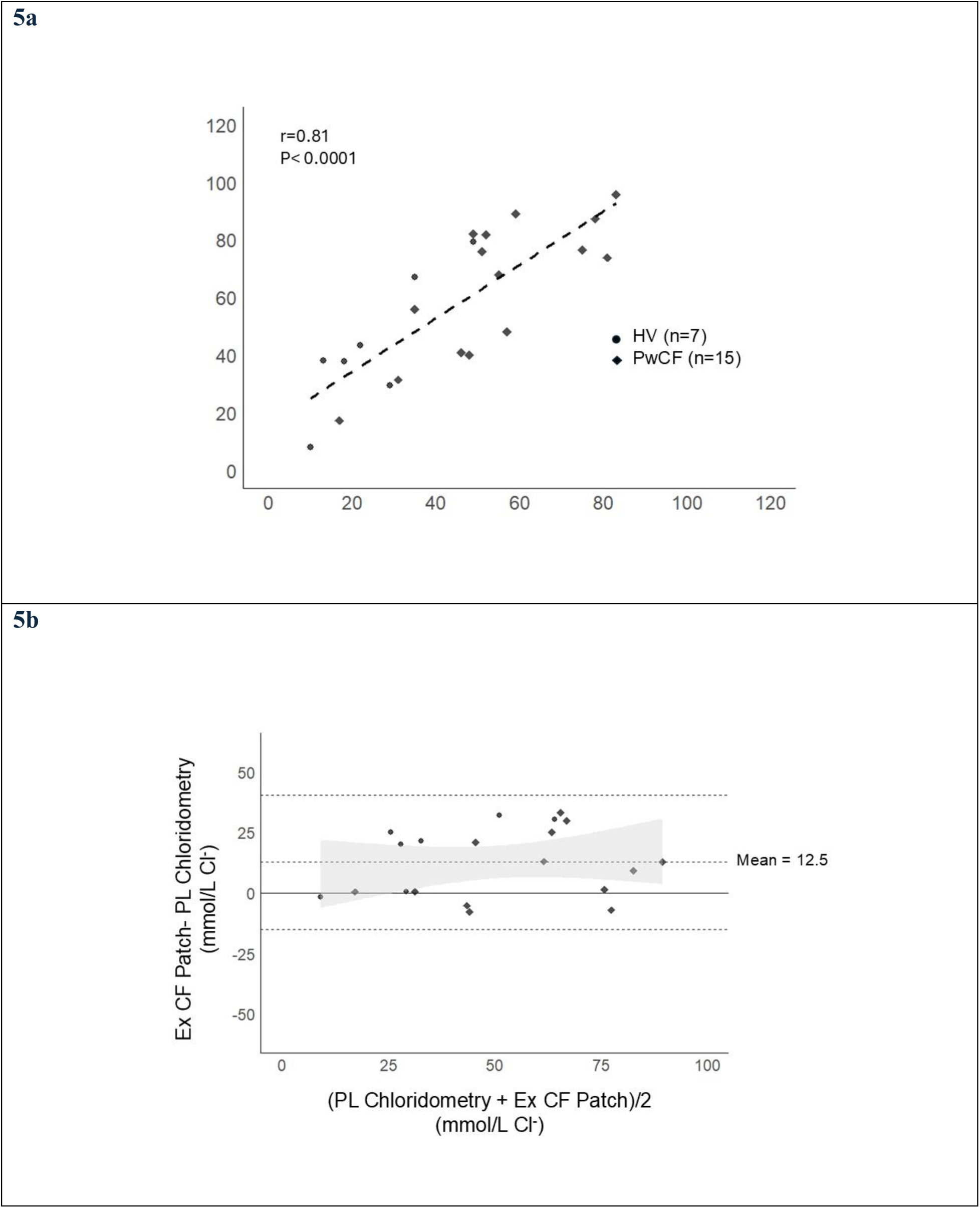
Exercise-induced CF Patch sweat chloride correlation versus pilocarpine-induced chloridometry sweat chloride. People with CF and HV had sweat collected with the CF patch during constant work exercise and it was compared to Pilocarpine induced sweat chloride measured by MSCS chloridometry. Shown is the regression line for the comparison of sweat chlorides between pilocarpine and exercise for the combined CF and HV cohort (a). The Bland-Alman plot for assessing agreement between the CF Patch and chloridometry shows agreement and a small bias. The dashed line shows the mean difference (bias), and the gray area represents the limits of agreement (mean difference ± 1.96 standard deviations) (b). When examined as separate cohorts HV and PwCF show similar correlations (c and d).

### Feasibility and Accuracy of Remote Exercise Sweat Chloride Collection

When evaluating success of remote exercise CF Patch use, we assessed whether participants were able to complete at least two interpretable CF Patch sweat collections. Of the 19 PwCF that started the remote exercise part of the study, all but 1 PwCF completed all 5 workouts as instructed. Of those, 15 (80%) PwCF reached the threshold of 2 interpretable CF sweat collections. Amongst the HV, all 7 completed the requested five collections (Figure 6a). For the successful remote sessions, average sweat [Cl^-^] across the successful remote exercise sessions was 56 (range 11 – 115 mmol/L) for PwCF and 31 (range 0.5 – 58 mmol/L) for HV. For the combined cohort there was a lower correlation between the mean remote exercise CF Patch sweat chloride and the exercise laboratory CF Patch sweat chloride, r=0.72 (*p*=0.0001) (Figure 6b). To ascertain the potential impact of CFTR modulators on the lower correlation between remote and laboratory exercise sweat chlorides, we analyzed HV and PwCF separately. In HV the correlation between remote and laboratory exercise sweat chlorides was excellent, r= 0.91 (*p*=0.004) (Figure 6c) but in PwCF there was a weaker correlation, r= 0.59 (*p*=0.02) (Figure 6d). This contrasts with the comparison between exercise laboratory CF Patch and pilocarpine-chloridometry sweat [Cl^-^], which showed that when analyzed separately, HV and PwCF had very similar correlations (Supplementary Figure 5a and 5b). Since pilocarpine-chloridometry and exercise laboratory CF Patch sweat [Cl^-^] were measured on the same day, this is consistent with the notion that CFTR modulators are contributing to more day-to-day variability of sweat [Cl^-^] in PwCF.

**Figure 6.**
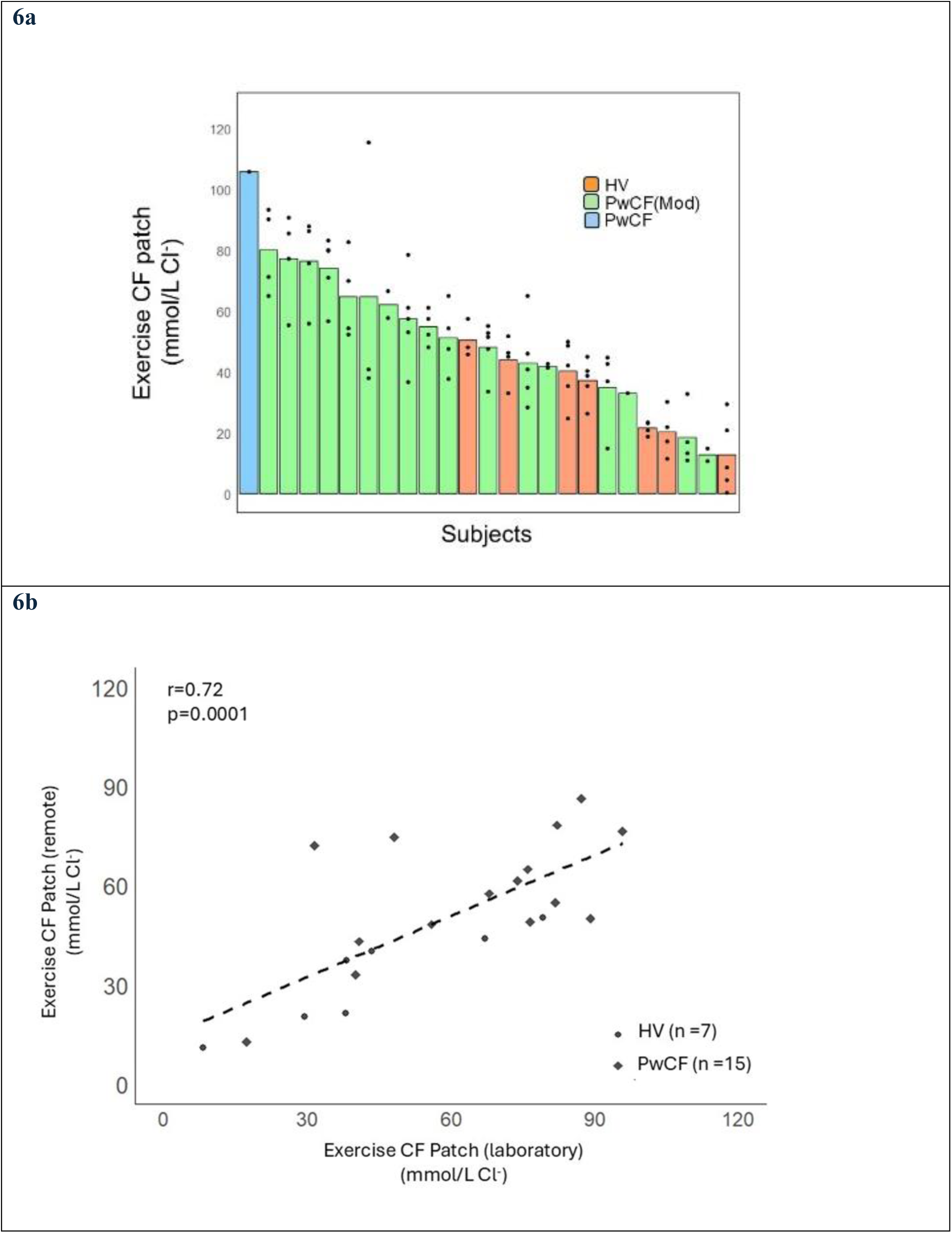
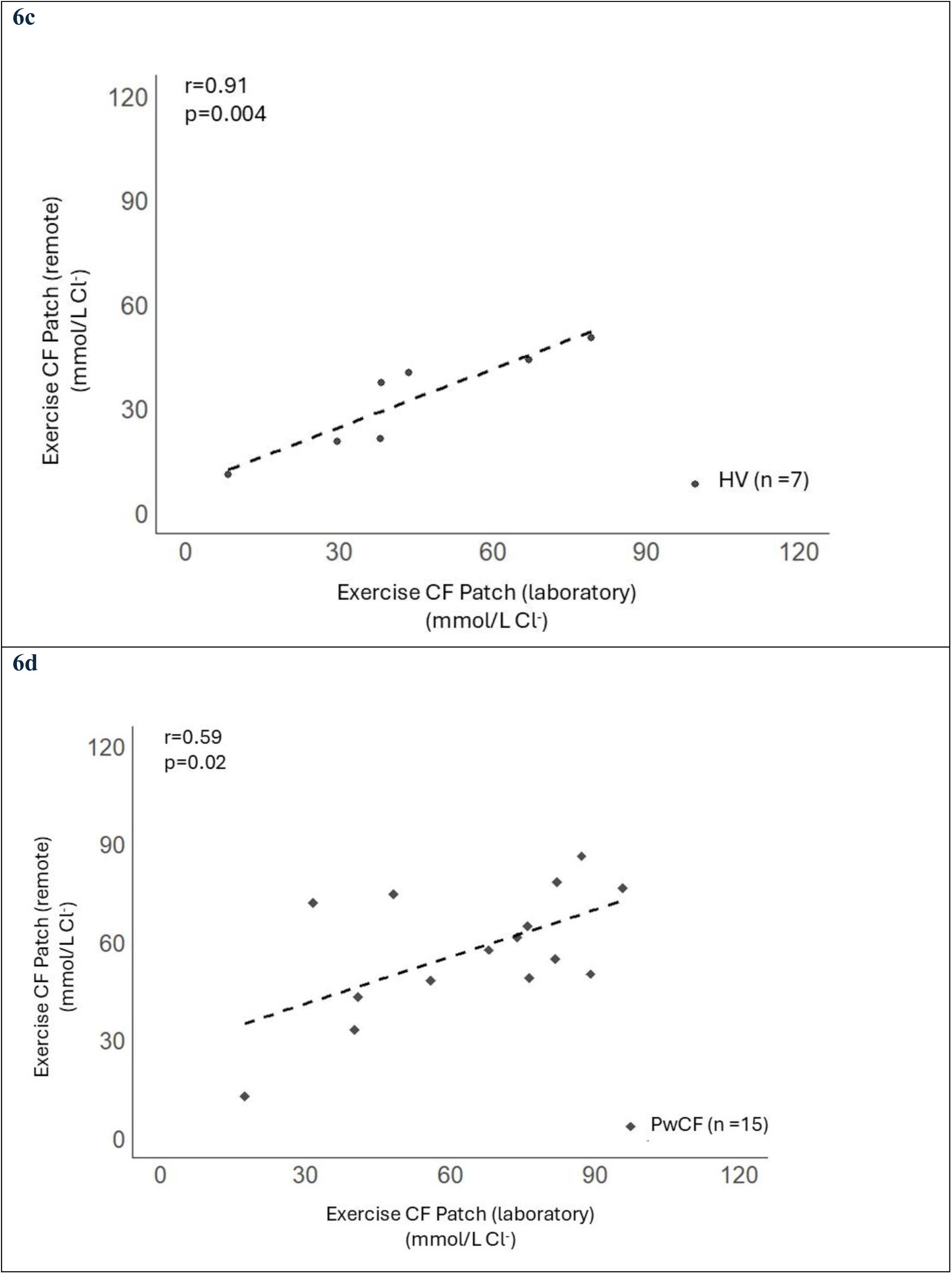
Remote exercise-induced CF Patch sweat chlorides have stronger correlation with exercise laboratory CF sweat chloride in HV than in PwCF. PwCF and HV had sweat chloride assessed sweat during remote exercise sessions and the mean of these values (a) for each individual was compared to the exercise laboratory sweat chloride collected during constant work exercise. Shown is the regression line for the comparison sweat chlorides between remote and laboratory exercise for the combined CF and HV cohort (b) and correlation was lower than the laboratory only comparisons. When examined as separate cohorts HV had a much stronger correlation than PwCF (c and d).

### Intra-Individual Variability in Sweat Chloride Concentrations

The difference between HV and PwCF in remote and laboratory exercise sweat chloride correlations prompted us to assess the intra-individual variability in remote exercise sweat chlorides in each cohort. We compared the standard deviations in remote exercise sweat chlorides between HV and PwCF. The average of the standard deviations for sweat chlorides in HV and PwCF was 15.1 and 24.2 mmol/L respectively, and differences in the standard deviations was statistically significant (*p*=0.03) (Figure 7). We then assessed the relationship between time of CFTR modulator intake and remote exercise CF Patch sweat [Cl^-^] for PwCF and found no significant correlation (data not shown). Similarly, there was no correlation between the volume of sweat collected during an individual exercise session and [Cl^-^] among HV or PwCF (data not shown). In supplementary table 1 we show the 4 sweat chloride values (single values for pilocarpine MSCS, pilocarpine CF Patch and laboratory exercise CF Patch and means for remote exercise CF Patch) collected for each subject enrolled in the study.

**Figure 7.**
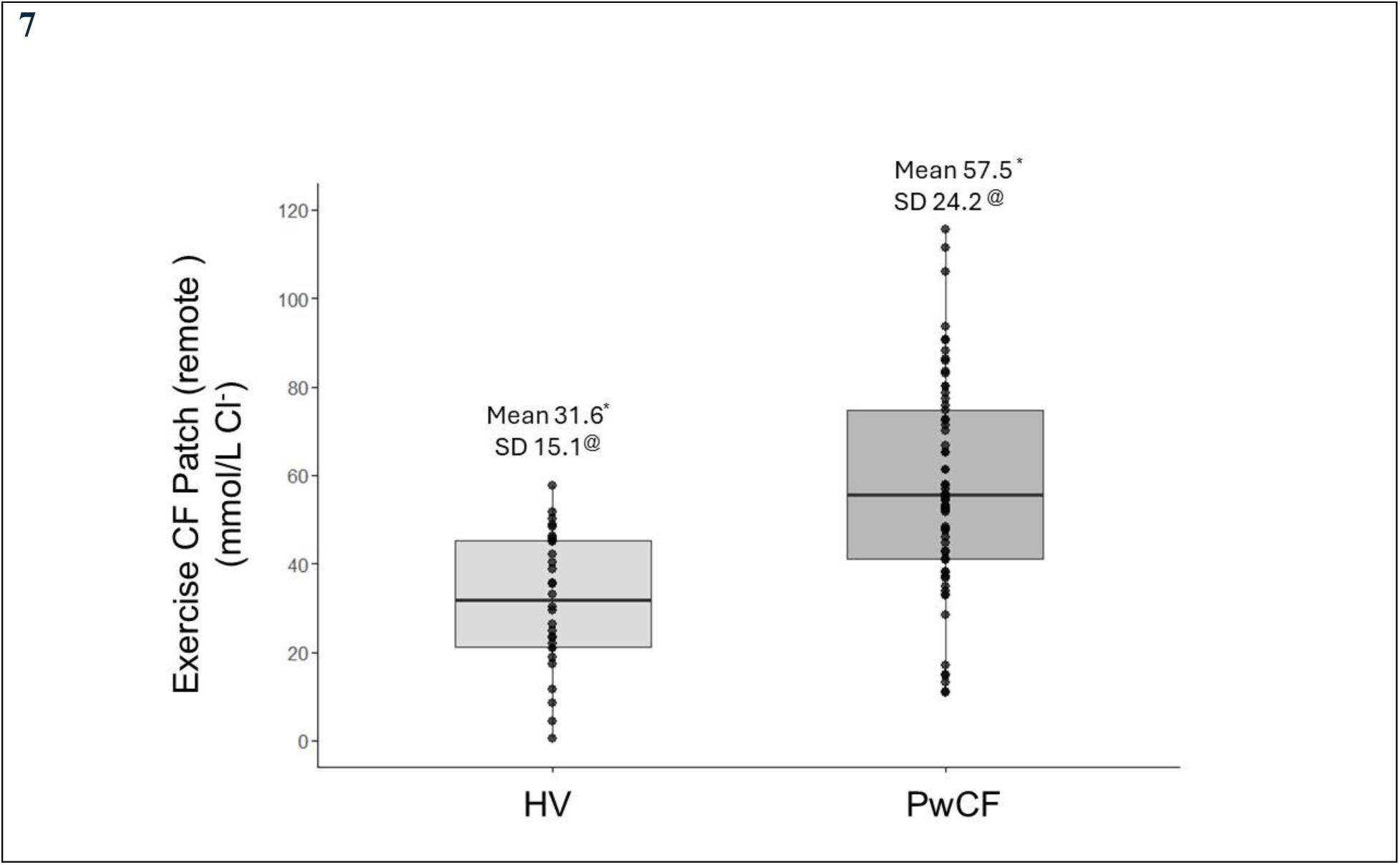
PwCF have greater variability in serial sweat chloride concentrations compared to HV by CF Patch. Each point represents the mean remote exercise sweat chlorides measured by CF Patch for HV and PwCF. We also calculated standard deviations within individuals for remote exercise sweat chlorides and compared HV and PwCF. PwCF had higher mean sweat chlorides and greater standard deviations (*p=0.01, ^@^p=0.03).

## Discussion

In this study we report four novel findings. We confirm for the first time that a microfluidic sweat patch using a colorimetric assay has a very strong correlation with MSCS chloridometry, the gold standard, in PwCF on CFTR modulators to measure reductions in sweat [Cl^-^]. In addition, we show a strong correlation between pilocarpine and exercise-induced sweat chlorides and demonstrate that serial remote sweat chloride measurement is feasible. In HV there is strong correlation between remote and laboratory exercise sweat chlorides, but this correlation is weaker in PwCF on CFTR modulators. Finally, our data suggest that some PwCF on CFTR modulators show substantial variability in sweat chlorides over a 2-week period.

These results further previous work, which demonstrated that a CF sweat patch and colorimetry can serve as an adequate surrogate for MSCS chloridometry and shows for the first time that it can detect CFTR modulator mediated decreases in sweat [Cl^-^].^9^ Importantly the CF Patch is accurate across a range of clinically relevant sweat [Cl^-^] values and did not demonstrate any systematic bias. The current CF sweat patch has design enhancements over previous iterations that allow for short collection periods (exercise time as low as 25 minutes), increased collection area for rapid sweat collection, and optimized color analysis in the Cl^-^ channel to function across a wider range of lighting conditions. Thus, the CF Patch offers several advantages over laboratory based chloridometry, such as portability, ease of repeat measurements and provides the possibility of measuring sweat [Cl^-^] in remote real-world settings including clinics or hospitals without access to a sweat lab and in low- and middle-income countries.^15^

Previous data have shown that in HV, exercise- and pilocarpine-induced sweat chlorides are similar at moderate intensity exercise,^16^ but there are no previous data on PwCF. Our data confirm that exercise is a satisfactory sweat inducer at higher sweat [Cl^-^] seen in PwCF and that exercise-induced sweat [Cl^-^] can serve as an adequate surrogate to pilocarpine sweat induction. Thus, exercise (or other ways to induce sweat) could be used for serial remote sweat [Cl^-^] measurements and these data could be utilized to inform multiple clinical aspects of care for PwCF. This includes but is not limited to assessing CFTR modulator adherence, impact of drug-drug interactions, pharmacodynamic effects of reduced CFTR modulator dosing or modest clinical responses to CFTR modulators. The data from the CF Patch could also be used to provide individualized recommendations for fluid and electrolyte repletion following exercise.^12^

The high success rate of remote sweat collection in PwCF suggests that it is feasible for patients to learn and use the CF Patch adequately. There was a lower percentage of successful collections in PwCF than HV due to the development of an intercurrent illness in 2 PwCF which is inherent in people with a chronic illness. Nevertheless, a large majority of PwCF were able to complete the threshold of 2 successful remote collections and it is likely that those who could not during the study period could do so once they recovered from their acute illness. This suggests that virtually all PwCF who exercise could successfully utilize the CF Patch.

Of note, compared to HV, PwCF had lower correlations between sweat [Cl^-^] measured in the exercise laboratory and those measured during remote exercise. For HV, intra-individual variations in sweat sodium concentration (which is highly correlated to sweat [Cl^-^]) is on the order of 5-16%, comparable to the present results.^17, 18^ PwCF showed a greater day to day variability of sweat [Cl^-^] compared to HV. Since all PwCF except 1 were on a CFTR modulator, we speculate that this may have been due to variable effects of modulators, though we cannot exclude the possibility that PwCF generally have more day-to-day variability in sweat [Cl^-^]. Large variations in repeated tests of sweat [Cl^-^] in PwCF have been observed previously, although it is unclear what fraction of participants in that study were on CFTR modulators.^19^ Other potential contributors to day-to-day variability include hydration status, salt intake or exercise intensity.^20^ Nevertheless, these data suggest that a single in clinic sweat [Cl^-^] measurement may not be an adequate surrogate for assessing chronic CFTR modulator pharmacodynamics in PwCF.

Previous data have shown limited correlation between single, clinic measured CFTR modulator-induced decreases in sweat [Cl^-^] and clinical responses.^3, 21–23^ It is possible that serial remote sweat [Cl^-^] measurements may offer a more accurate assessment of individual CFTR modulator exposure and a better correlation with clinical response. This study demonstrates the potential for integrating exercise-induced sweat [Cl^-^] testing into clinical monitoring protocols for PwCF on CFTR modulators. The CF sweat patch could provide a less invasive, patient-friendly alternative for longitudinal monitoring, particularly for assessing CFTR modulator efficacy and side effects, identifying potential causes of variability in therapeutic responses and facilitating research studies in non-clinical settings. This could be studied in a prospective fashion to provide evidence for clinical utility before implementing into regular clinical use.

## Methods

### CF Patch device design and fabrication

A detailed description of the CF Patch and smartphone application has been published previously.^12, 13^ Briefly, a multi-layered stack of thin-film polymeric materials (polyurethane film; Innovize Inc., MN, USA) with laser-cut regions creates the micro-channels and inlet ports. Pressure-sensitive adhesive bonds the intermediate fluidic layer with the top graphics and subjacent skin adhesive layers (polyethylene film; 3M, MN, USA). Laser-cut inlet windows in the adhesive layer create openings to define the two sweat collection regions for microchannel 1 and microchannel 2. For colorimetric analysis of microchannel 1 (volume of approximately 112 µL), a dehydrated colored dye (≈4 µL) was deposited in an assay well region and used to mix with sweat, providing an optical method to measure regional sweat volume and sweating rate. Microchannel 2 (volume approximately 20 µL) was embedded with a mixture of silver chloranilate (Green Room Board Co, NC, USA) and polyhydroxyethylmethacrylate (pHEMA; Sigma-Aldrich, MO, USA) hydrogel placed near the assay well region to mix with sweat for Cl concentration analysis.^12^ Images of the CF Patch were captured using the CF application (beta) for colorimetric analysis. Images were initially stored on the local smart phone memory, then transmitted to a computer via the cloud services operated by Apple Inc.

### Study design

We conducted a prospective, single-center, feasibility study of adult PwCF and adult healthy volunteers (HV) who were self-identified as exercising at home. The study was approved by the Northwestern University Biomedical Institutional Review Board (IRB) (STU00215214). Participants were recruited between April 2022 and May 2024. PwCF were recruited from a single CF center at Canning Thoracic Institute (Northwestern Medicine) and HV were recruited from the community. All participants were informed of the experimental procedures and associated risks before providing consent.

Each participant was asked to complete 2 parts of the study. Participants first completed clinic-based sweat testing, consisting of resting pilocarpine-induced sweat collection, a cardiopulmonary exercise test (CPET) and on-site exercise-induced sweat collection at the exercise physiology laboratory of Department of Physical Therapy & Human Movement Sciences (Northwestern University). After completing the on-site portion, participants were asked to complete 5 remote exercise sessions over 14 days while wearing the CF sweat patch. The location and activity of remote exercise were of their own preference.

A total of 23 PwCF and 7 healthy volunteers (HV) consented to participate in this study. Prior to the on-site visit, 3 PwCF withdrew consent and were excluded from analyses. One PwCF experienced a pulmonary exacerbation after completing 3 remote exercise sessions and all data collected prior to early termination were included in analyses.

### Human subject sweat testing

Our first objective was to collect pilocarpine-induced sweat and compare MSCS collection and chloridometry with the CF Patch colorimetric analysis for baseline sweat [Cl^-^]. Sweat was induced via pilocarpine iontophoresis on the left and right ventral forearms and collected into a CF Patch and Macroduct coil in separate arms. Samples extracted from the Macroduct coil were analyzed at the CF Therapeutics Development Network (TDN) Center for Sweat Analysis at Children’s Hospital Colorado.

CPET consisted of an incremental protocol on a cycle ergometer (LODE, The Netherlands). Once initiated, work rate increased incrementally every minute until the subject was no longer able to maintain a cycling cadence of ≥ 60 revolutions per minute. Rating of perceived exertion and exercising blood pressure values were recorded every two minutes during the CPET. VO_2peak_ was determined using 30-second averaging, and the highest single 30-second value was recorded as VO_2peak_. This value was recorded during the last minute of exercise. Exercising heart rate data throughout the study were monitored by a Medtronic Zephyr biomodule and chest strap (Medtronic, Minneapolis, MN).

Our second objective was to collect exercise-induced sweat into the CF sweat patch to measure regional sweat volume and [Cl^-^]. After completion of the CPET, participants were allowed a dedicated rest period and fluid replenishment (water and Gatorade) until their baseline heart rate was reached and post-CPET sweat induction had ceased. Research personnel then applied two microfluidic sweat patches, one on each arm following which each subject began exercising at a constant wattage which was 50-60% of peak wattage derived from the CPET. Exercising heart rate and rating of perceived exertion, as well as environmental conditions including temperature, barometric pressure, and humidity of the exercise physiology laboratory were recorded every 5 minutes during on-site exercise. Participants continued exercising until either there was sufficient sweat in each channel of the CF Patch (assessed by visual inspection) or 60 minutes had elapsed.

### Remote exercise sessions and sweat collection

Following recovery from exercise testing, participants were provided with CF sweat patches and a Medtronic Zephyr biomodule with chest strap to wear during the 5 remote exercise sessions over a 14-day period. Subjects were trained on use of all devices prior to starting remote workouts. They were also provided with a smart phone with the OmniSense mobile application. There were no specific requirements for the remote exercise sessions and participants were instructed to perform their normal exercise activities (e.g., running/cycling, resistance training, hot yoga). Participants were instructed to take photos of the CF sweat patches following each of the 5 remote exercise sessions and transmit the images to the study team which were subsequently analyzed.

### Data collection

Study data were collected and managed using Research Electronic Data Capture (REDCap) tools hosted at Northwestern University.^24–26^ REDCap is a secure, web-based software platform designed to support data capture for research studies, providing 1) an intuitive interface for validated data capture; 2) audit trails for tracking data manipulation and export procedures; 3) automated export procedures for seamless data downloads to common statistical packages; and 4) procedures for data integration and interoperability with external sources.

### Smartphone and camera-based colorimetric analysis of sweat volume and Cl concentration

Details on smartphone colorimetric analysis under controlled and ambient lighting conditions have been published.^11^ Briefly, photos of microfluidic patches were captured using a smartphone application (iPhone 11, Apple Inc.) in clinical and remote settings. Images were captured at the time of CF Patch removal, at the end of sweat-induction by pilocarpine or exercise. When captured in a clinical setting multiple images were captured by study personnel within 5 minutes of completion of sweat collection. At home, study participants were instructed to capture multiple CF images within 5 minutes of exercise completion. To assess feasibility of remote image capture, success was deemed if a participant recorded interpretable images from at least 2 remote exercise sessions. Final mean volume and sweat [Cl^-^] were calculated from manual image analysis. Sweat [Cl^-^] and volume data from CF Patch collections were obtained from each sample from which at least a single image measurement was available. We also performed an additional analysis in which we only included data from which triplicate images were obtained which is shown in supplementary data (Supplementary Figures 1-4). RAW images were used for processing to eliminate artifacts introduced by normalization, compression, and other preprocessing steps. The sweat volume and [Cl^-^] were calculated by manual image analysis by a trained data scientist.^11^ The colors of four swatches on the CF Patch (green, purple, orange, white) were measured and used to correct for variations in white balance due to ambient lighting. Local sweat volume was measured by identifying the furthest extent to which colored dye propagated in microchannel 1 and mapping that location to a volume using a CAD model of the patch. Chloride was measured by computing the average color within microchannel 2 relative to the background color of the patch. In CIELAB color space this color is represented by a 2D vector. This vector was then projected onto the unit vector for the color of chloranilic acid, which was measured empirically. The length of this projection maps to chloride concentration by a power law relationship, as described by the Beer-Lambert law.

### Statistical analysis

Statistical analysis was performed using the R statistical language version 4.4.2. Descriptive statistics are reported as means and standard deviations. Comparisons between HV and PwCF and between different conditions were analyzed using independent samples t-tests. Bivariate comparisons between measurement conditions (i.e. MSCS vs. CF patch etc.) were made by linear regression and Pearson correlation coefficients were calculated. Proportional bias was calculated for each Bland-Altman analysis. Levene’s test was used for comparison of variance measurements of remote exercise sweat chlorides. Statistical significance was attributed when *p* < 0.05.

## Data availability

All data needed to evaluate the conclusions in the paper are present in the paper and/or the Supplementary Materials. Data sets generated during the current study are available from the corresponding authors on reasonable request.

## Code availability

Portions of software code that are central to interpreting or replicating the current study are available from the corresponding author on reasonable request.

## Acknowledgements

We thank Dr. Susanna McColley and Gabriel Fagans for their support in defining key sensing features and fabrication of CF Patches. This work made use of the Northwestern University’s Querrey Simpson Institute for Bioelectronics for testing and analysis. Research reported in this publication was supported, in part, by the National Institutes of Health’s National Center for Advancing Translational Sciences (Grant Number UL1TR001422) the National Heart, Lung, and Blood Institute Grant Number (R43HL162246). The content in this manuscript is solely the responsibility of the authors and does not necessarily represent the official views of the National Institutes of Health.

## Author contributions

R.N. and M.J. conceived the idea, designed the research, did the data analysis and wrote the manuscript. A.J.A., R.G., J.A.R., J.A.W., D.S.Y. conceived the idea, designed the research and wrote the manuscript. T.C. and J.S. did the data analysis and wrote the manuscript. M.G., Z.K., M.C. and A.D. implemented the study protocol. G.J.G. and D.M.C. helped design the protocol and write the manuscript. A.J.A., D.E.W., J.C.W. performed and were involved in the manufacturing of the sensors. A.J.A., J.C.W., D.E.W., performed software design and software validation. D.E.W., J.C.W., A.J.A. assisted in device fabrication and field testing.

## Competing interests

A.J.A., R.G., and J.A.R. are co-founders of Epicore Biosystems, which develops and commercializes microfluidic devices for sweat analysis. D.E.W., J.C.W., and J.A.W. are currently employed by Epicore Biosystems. All other authors declare that they have no competing interests.

## Notes

### Author Declarations

The study was approved by the Northwestern University Biomedical Institutional Review Board (IRB) (STU00215214).

