## Supplementary Data for "Remote exercise-induced sweat chloride measurements using a wearable microfluidic sticker in cystic fibrosis patients"

**Supplementary Figure 4| Remote exercise induced CF patch sweat chlorides correlation with exercise laboratory CF sweat chloride.** PwCF and HV had sweat chloride assessed sweat during remote exercise sessions and the mean of these values for each individual was compared to the exercise laboratory sweat chloride collected during constant work exercise. Shown is the regression line for the comparison sweat chlorides between remote and laboratory exercise for the combined CF/ HV cohort (a) and correlation was lower than the laboratory only comparisons.

**Supplementary Figure 5 | Exercise induced CF Patch sweat chloride has similar correlation with Pilocarpine induced chloridometry in HV and PwCF**. PwCF and HV had sweat collected with the CF Patch during constant work exercise and it was compared to Pilocarpine induced chloridometry. Shown is the regression line for the comparison sweat chlorides between pilocarpine and exercise for individual HV (a) and PwCF (b).

**Supplementary Table 1 | 4 sweat chloride concentration measurements for each study participant.**

### Supplementary Figures

**SF1a**

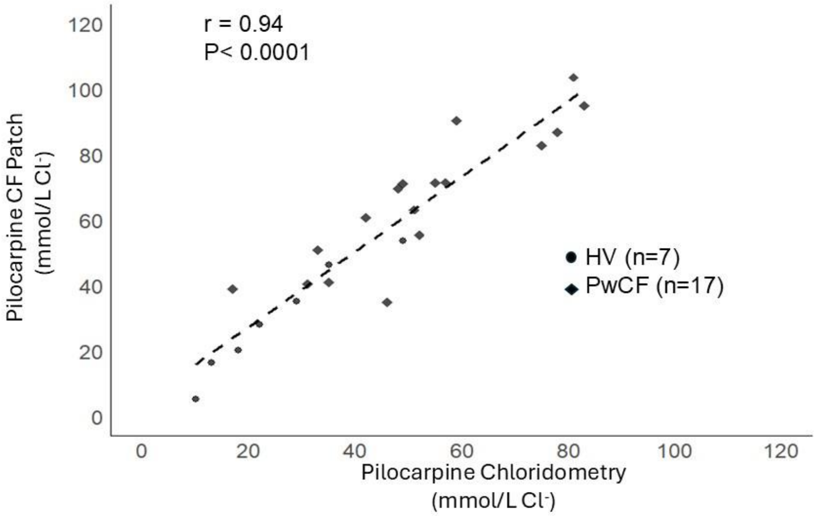


**SF1b**

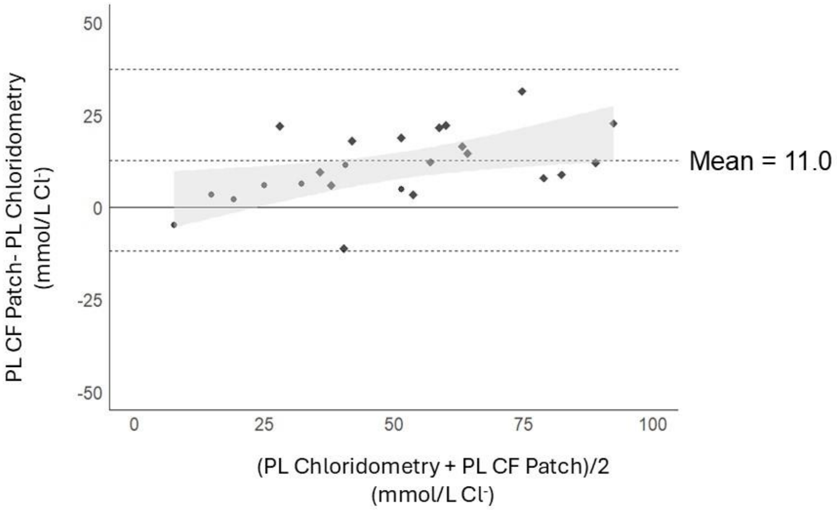

**SF2a**

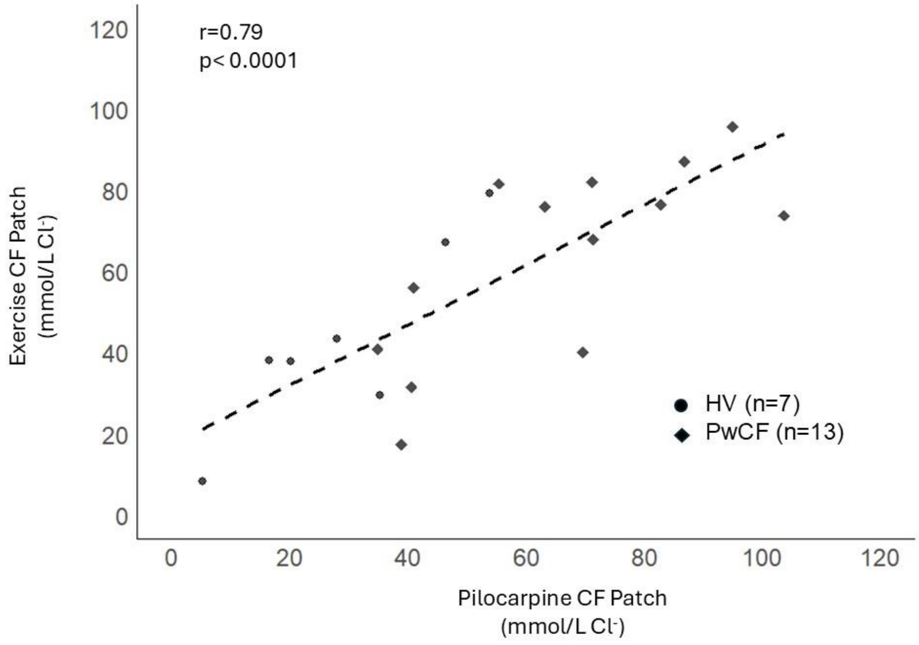
r=0.79
p< 0.0001
120
100
80
60
40
20
0
Exercise CF Patch (mmol/L Cl-)
0
20
40
60
80
100
120
Pilocarpine CF Patch (mmol/L Cl-)
HV (n=7)
PwCF (n=13)

**SF2b**

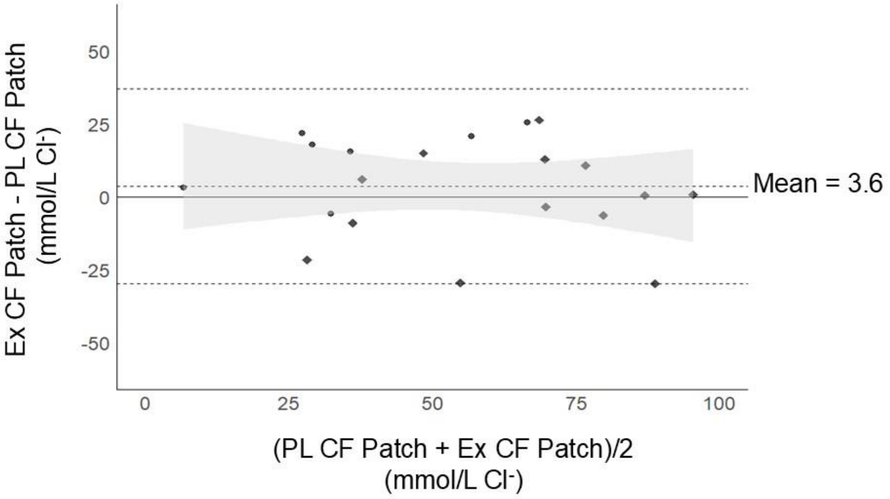
50
25
0
-25
-50
Ex CF Patch - PL CF Patch (mmol/L Cl-)
Mean = 3.6
0
25
50
75
100
(PL CF Patch + Ex CF Patch)/2 (mmol/L Cl-)

**SF3a**

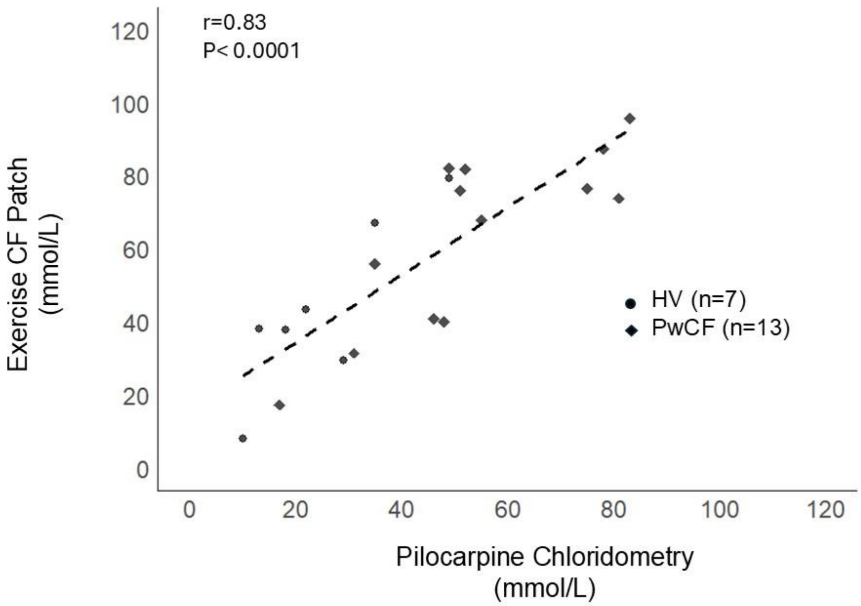
r=0.83
P< 0.0001
Exercise CF Patch
(mmol/L)
120
100
80
60
40
20
0
0
20
40
60
80
100
120
HV (n=7)
PwCF (n=13)
Pilocarpine Chloridometry
(mmol/L)

**SF3b**

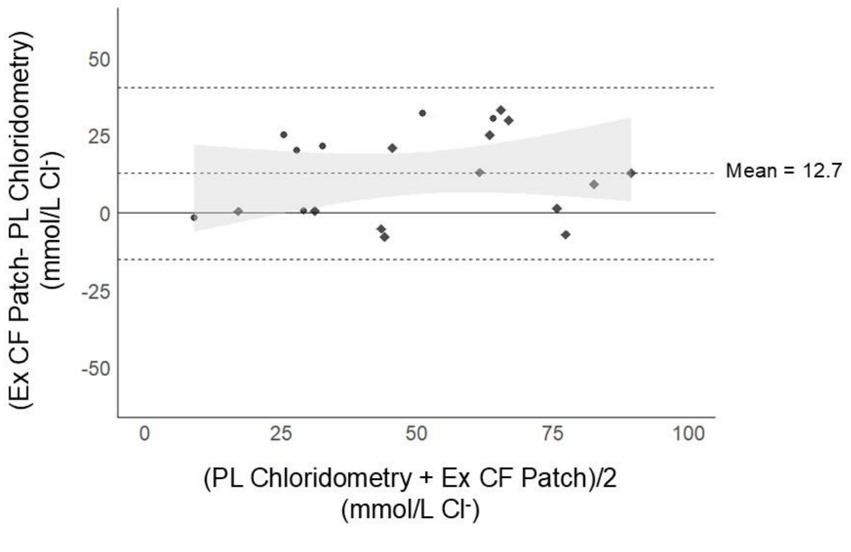
(Ex CF Patch- PL Chloridometry)
(mmol/L Cl-)
50
25
0
-25
-50
0
25
50
75
100
Mean = 12.7
(PL Chloridometry + Ex CF Patch)/2
(mmol/L Cl-)

**SF4**

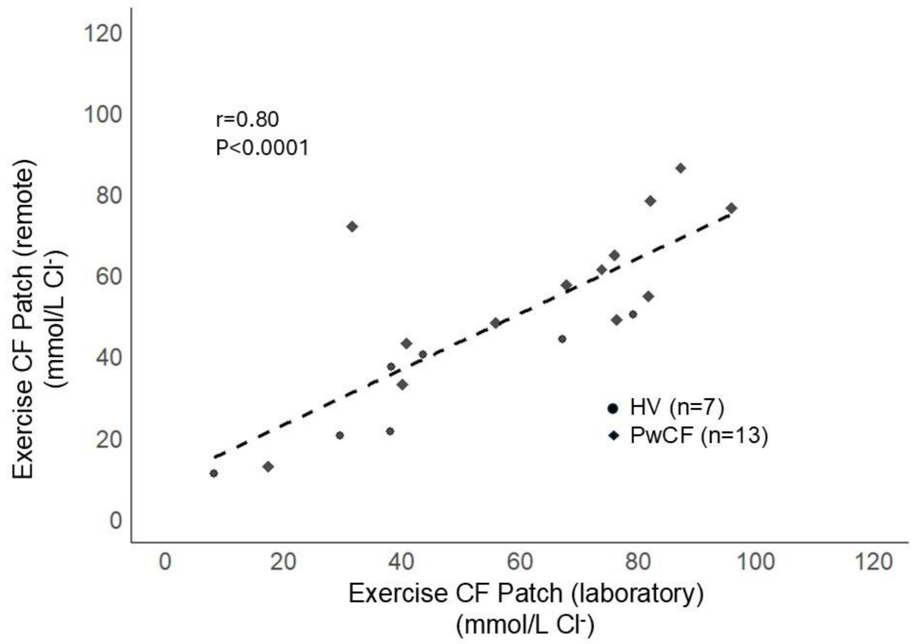

r=0.80
P<0.0001
Exercise CF Patch (remote) (mmol/L Cl-)
Exercise CF Patch (laboratory) (mmol/L Cl-)
HV (n=7)
PwCF (n=13)
0
20
40
60
80
100
120

**SF5a**

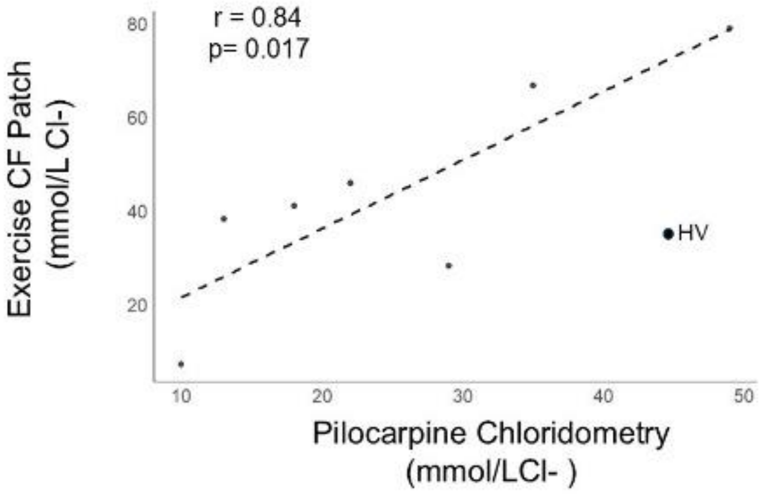

r = 0.84
p= 0.017
Exercise CF Patch (mmol/L Cl-)
80
60
40
20
10
20
30
40
50
HV
Pilocarpine Chloridometry (mmol/LCl- )


**SF5b**

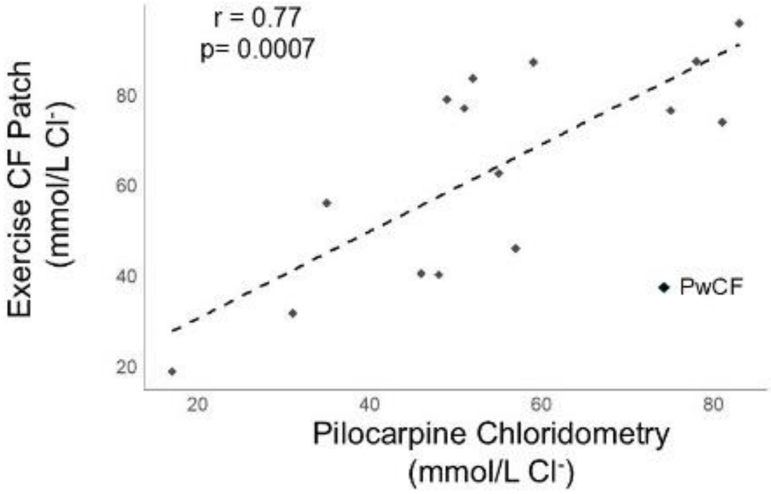

r = 0.77
p= 0.0007
Exercise CF Patch (mmol/L Cl-)
80
60
40
20
20
40
60
80
PwCF
Pilocarpine Chloridometry (mmol/L Cl-)

| ID | MSCS | Pilo Patch | Lab Patch | Remote Patch |
|---|---|---|---|---|
| HV 1 | 10 | 9 | 7 | 11 (0 - 30) |
| HV 2 | 18 | 22 | 43 | 21 (19 - 26) |
| HV 3 | 35 | 48 | 63 | 44 (33 - 56) |
| HV 4 | 13 | 17 | 38 | 37 (24 - 49) |
| HV 5 | 22 | 26 | 43 | 40 (23 - 51) |
| HV 6 | 29 | 36 | 28 | 20 (8 - 32) |
| HV 7 | 49 | UN | 79 | 50 (43 - 60) |
| PwCF 1 | 51 | 63 | 75 | 65 (47 - 85) |
| PwCF 2 | 17 | 38 | 20 | 13 (10 - 17) |
| PwCF 3 | 46 | 35 | 39 | 43 (28 - 65) |
| PwCF 4 | 57 | 75 | 46 | 75 (54 - 93) |
| PwCF 5 | 55 | 68 | 62 | 58 (35 - 85) |
| PwCF 6 | 106 | UN | UN | 106 (99 - 113) |
| PwCF 7 | 54 | 106 | UN | UN |
| PwCF 8 | 49 | 75 | 74 | 78 (51 - 95) |
| PwCF 9 | 31 | 40 | 31 | 72 (38 - 118) |
| PwCF 10 | 81 | 97 | 70 | 61 (53 - 68) |
| PwCF 11 | 52 | UN | 87 | 55 (47 - 62) |
| PwCF 12 | 35 | 44 | 55 | 48 (30 - 58) |
| PwCF 13 | 33 | 52 | UN | 19 (9 - 36) |
| PwCF 14 | 83 | 90 | 90 | 77 (53 - 93) |
| PwCF 15 | 59 | 88 | 87 | 50 (40 - 73) |
| PwCF 16 | 42 | 53 | UN | 35 (13 - 46) |
| PwCF 17 | 75 | 81 | 77 | 49 (35 - 68) |
| PwCF 18 | 48 | 69 | 39 | 33 (29 - 36) |
| PwCF 19 | 78 | 87 | 90 | 86 (61 - 114) |

**Supplementary Table 1**
